# Protocol for secondary data analysis of 4 UK cohorts examining youth adversity and mental health in the context of intersectionality

**DOI:** 10.1101/2023.07.19.23292906

**Authors:** Georgina M. Hosang, Laura Havers, Ruichong Shuai, Peter Fonagy, Mina Fazel, Craig Morgan, Alexis Karamanos, Daisy Fancourt, Paul McCrone, Melanie Smuk, Kamaldeep Bhui, Sania Shakoor

**Affiliations:** Centre for Psychiatry and Mental Health, Wolfson Institute of Population Health, Queen Mary, University of London, London, UK; Anna Freud National Centre for Children and Families, London, UK; Research Department of Clinical, Educational and Health Psychology, University College London, London, UK; Department of Psychiatry, University of Oxford, Oxford, UK; Health Service and Population Research, Institute of Psychology, Psychiatry & Neuroscience, King’s College London, London, UK; Centre for Society and Mental Health, King’s College London, London, UK; Department of Population Health Sciences, King’s College London, London, UK; Department of Behavioural Science and Health, University College London, London, UK; Institute for Lifecourse Development, University of Greenwich, London, UK; Centre for Genomics and Child Health, Blizard Institute, Queen Mary, University of London, London, UK; Department of Psychiatry, Nuffield Department of Primary Care Health Sciences and Wadham College, University of Oxford, Oxford, UK; Oxford Health and East London NHS Foundation Trusts, UK; World Psychiatric Association Collaborating Centre, Oxford, UK

**Keywords:** Adverse Childhood Experiences, Adversity, Depression, Anxiety, Youth Mental Health, Intersectionality, Ethnicity, Gender, Socio-Economic Status, Neurodivergence

## Abstract

**Background:** Youth adversity (e.g., abuse and bullying victimisation) is robust risk factor for later mental health problems (e.g., depression and anxiety). Research shows the prevalence of youth adversity and rates of mental health problems vary by individual characteristics, identity or social groups (e.g., gender and ethnicity). However, little is known about whether the impact of youth adversity on mental health problems differ across the intersections of these characteristics (e.g., white female). This paper reports on a component of the ATTUNE research programme (work package 2) which aims to investigate the impact and mechanisms of youth adversity on depressive and anxiety symptoms in young people by intersectionality profiles.

**Methods:** The data are from 4 UK adolescent cohorts: HeadStart Cornwall, Oxwell, REACH, and DASH. These cohorts were assembled for adolescents living in distinct geographical locations representing coastal, suburban and urban places in the UK. Youth adversity was assessed using a series of self-report questionnaires and official records. Validated self-report instruments measured depressive and anxiety symptoms. A range of different variables were classified as possible social and cognitive mechanisms.

**Results and analysis:** Structural equation modelling (e.g., multiple group models, latent growth models) and multilevel modelling will be used, with adaptation of methods to suit the specific available data, in accord with statistical and epidemiological conventions.

**Discussion:** The results from this research programme will broaden our understanding of the association between youth adversity and mental health, including new information about intersectionality and related mechanisms in young people in the UK. The findings will inform future research, clinical guidance, and policy to protect and promote the mental health of those most vulnerable to the negative consequences of youth adversity.

## Introduction

Mental health problems, such as depression (e.g., low mood and irritability) and anxiety (e.g., worry and feeling on edge) affect up to 17% of young people in the UK, a figure that has been steadily rising since 2004 [1]. In fact, up to 50% of mental health problems manifest by adolescence [2], highlighting the importance of clinical and research efforts focused on this developmental period. Youths experiencing depression and anxiety are more likely than their peers to experience social exclusion and discrimination, which in turn can amplify the risk of self-harm and suicide [3]. Against this background it is evident that tackling youth mental health is a public health concern.

The prevalence and consequences of mental health problems vary significantly by individual-level demographic characteristics, identities and groups (referred to hereon in as individual characteristics), such as ethnicity, gender, socio-economic status [SES] and (innate) neurodivergence (e.g., attention deficit hyperactivity disorder [ADHD]) [4–7]. For instance, the rates of depressive and anxiety symptoms have been found to be over twice as high for youths with low SES compared to those with higher SES [7]. Importantly, there is evidence to suggest that the *intersection* of such characteristics are associated with greater mental health problems (e.g., females from low SES backgrounds) [8] and as such, there is a need for these to be systematically investigated.

Youth adversity is a robust risk factor for later mental and physical health problems [9, 10]. Youth adversity can be defined as stressful and in some cases traumatic experiences that occur during childhood and/or adolescence [11]. Youth adversity is an umbrella term that covers traditionally defined “adverse childhood experiences” that are take place in the home (abuse, neglect, parental mental illness and exposure to domestic violence [12]) as well as those that can occur in other settings (e.g., bullying victimisation). Many of these experiences are potentially preventable, providing policy makers, health, education and social care practitioners opportunities for prevention and intervention [13, 14]. There is emerging evidence to suggest that the *prevalence* of youth adversity varies by individual-level demographic characteristics [4, 15-17] and their intersection [18]. However, little is known about the role of intersectionality in the *relationship* between youth adversity and mental health problems [19].

The prevalence of youth adversity and mental health problems varies by geographical locations or ‘place’ across the UK [22, 23]. The degree of urbanicity (urban to rural), for instance, has been linked to depression [24]. Place is important because it can create circumstances and contexts of multiple disadvantage, from structural (e.g., access to health care), community (e.g., crime) to personal levels [25]. Further investigation is needed to understand the magnitude of the association between youth adversity and mental health problems by place warrants further investigation. According to theories of intersectionality individual characteristics (e.g., gender, ethnicity, neurodivergence, SES) not only reflect positions within societal and social hierarchies but also that the intersection of these characteristics gives rise to unique social experiences in the context of oppression and privilege [20, 21]. To understand the extent and ways in which intersectionality profiles are associated with risk and resilience of psychopathology in the face of youth adversity, it is crucial to include large diverse samples.

There is a large body of research focused on the protective effects of social and cognitive factors in the relationship between youth adversity and mental health in young people. Factors, such as, social support (practical and emotional aid provided by friends and family) and attributional style (attribution and interpretation of one’s experiences) have been shown to buffer from depressive and anxiety symptoms in the face of youth adversity [16]. However, little is known about whether these effects vary between groups based on individual characteristics (e.g., gender) and the intersection of such (e.g., gender and SES). This area requires further research attention before any conclusions can be drawn.

In this paper, we present a protocol as part of work package 2 of the ATTUNE project, which aims to investigate youth adversity and mental health through an intersectionality lens, using existing data from community cohorts in different UK geographical locations.

### Research questions and hypotheses

ATTUNE is a multi-site study which aims to explore young people’s experiences and understandings of mental health and adversity using arts-based methods. In this protocol we present work package 2, which is designed to explore the role and mechanisms of youth adversity on mental health problems in young people by intersectionality profiles. Specifically, the work package will address the following 4 research questions (RQ).

RQ1. Does the *prevalence* of youth adversity, depressive and anxiety symptoms vary by place? ***Hypothesis:*** more youth adversity and depressive and anxiety symptoms will be observed in inner-city locations compared with suburban and coastal places.
RQ2. To what extent does the *relationship* between youth adversity and depressive and anxiety symptoms vary by intersectionality profiles? ***Hypothesis:*** The magnitude of the association between youth adversity and depressive and anxiety symptoms will be the greatest at the intersections of multiple disadvantaged social positions (e.g., females, high neurodivergence, low SES, from ethnic minority backgrounds).
RQ3. To what extent do the *developmental trajectories* of depressive and anxiety symptoms vary by youth adversity and intersectionality profiles? ***Hypothesis:*** Developmental trajectories will vary by youth adversity and intersectionality profiles. It is expected that youth adversity compared to no youth adversity will be associated with greater baseline as well as more stable/persisting depressive and anxiety symptoms over time, and that there will be some degree of moderation by intersectionality profile.
RQ4. To what extent is the relationship between youth adversity and depressive and anxiety symptoms *moderated by social support and social cognitive factors*? And does this vary by intersectionality profiles? ***Hypothesis***: the relationship between youth adversity and depressive and anxiety symptoms will be moderated by social support and social cognitive factors. No predictions are made regarding differences between intersectionality profiles.

## Methods

### Study design

Secondary statistical analysis of existing quantitative data drawn from cohort studies.

### Cohorts

The research questions outlined earlier for the research programme will be addressed using 4 UK cohorts, each of which will be described below and summarised in ***Table 1***.

***HeadStart Cornwall*** cohort consists of over 12,000 young people aged between 11-16 years from Cornwall, in the South West coast of England, UK [26]. This cohort has cross-sectional and longitudinal data available. Pupils attending all state schools in Cornwall were invited to participate, thus the sample size was not limited or predetermined. The study adopted a school-based and parental opt-out approach for participant recruitment. This reduces potential bias associated with opt-in recruitment approaches and recruitment reliant on advertisements in particular locations (physical and online). Year 7 pupils (aged between 11 and 12) at the first wave of assessment in 2017 were followed longitudinally through to Year 12 (ages 16 and 17). Data was collected across 2017-2022. Data linkage is available for this cohort with the National Pupil Database providing information about the child and their family’s background and receipt of free school meals.

***OxWell*** is an ongoing cross-sectional study, which had over 30,000 individuals that participated in 2021 [27]. Participants are aged between 8-18 years and have been recruited from over 180 schools and Further Education Colleges in England, UK (Berkshire, Buckinghamshire, Gloucestershire, Wiltshire, Bristol, South Somerset, and Liverpool) since 2019. All schools (primary and secondary) and Further Education Colleges within the target counties were invited to take part, and all pupils attending participating schools/colleges were invited to participate. Therefore, the sample size was not limited or predetermined. Participant recruitment was based on school-based and parental opt-out. For the purpose of this research programme, only data from individuals aged 13-18-years will be analysed.

***REACH [Resilience, Ethnicity and AdolesCent mental Health]*** is a longitudinal cohort study based in London, UK [28]. Participants were recruited from 12 secondary schools from two South London Boroughs, Lambeth and Southwark. The schools were selected to be representative of the 38 mainstream schools in these boroughs based on ethnicity and socio-economic status. Participants were recruited using a parental opt-out approach and were aged 11-14 years at baseline in 2015 and were followed up 1 and 2 years later in the first phase of the study. The cohort consists of over 4,000 young people. The sample size was determined by power analysis calculations, based on hypothesised effect sizes, and accounting for both attrition and inflation attributable to clustering within schools.

***DASH [Determinants of Adolescent Social well-being and Health] Study*** is a longitudinal cohort of over 6,500 youths aged between 11-13 years old at baseline recruited from 51 schools from across London, UK, in 2003 [29]. To be eligible to take part, participants had to be in Years 7 or 8 (aged between 11-13 years) attending a participating secondary school in the London Boroughs of Brent, Croydon, Hackney, Hammersmith & Fulham, Haringey, Lambeth, Newham, Southwark, Waltham Forest and Wandsworth. Boroughs and schools were specifically chosen to enable representation of individuals from ethnic minority groups from a range of academic performance standards. DASH adopted a school-based and parental opt-out approach to recruiting participants. The cohort was followed up when they were aged 13-15 years, and a pilot follow-up was conducted at age 21-23 years [30].

**Table 1.** Description of cohorts.

| Cohort | N | Location | Age range | Youth adversity measure | Depression and anxiety measures | Neurodivergence | SES | Mechanisms | Ethical approval |
| --- | --- | --- | --- | --- | --- | --- | --- | --- | --- |
| HeadStart Cornwall | Over 12,000 | Cornwall (coastal) | 11-16 years | 2 Scales:<br>1. Bullying victimisation SDQ item<br>2. Family List- range of experience including<br>• Child at risk of sexual exploitation<br>• Family homelessness<br>• Exposure to domestic violence<br>• Unmanaged physical or mental illness in the household | SDQ emotional problems subscale | SDQ hyperactivity/inattention subscale | Receipt of free school meals | - | University College London Ethics Committee (ref: 8097/003) |
| OxWell | Over 30,000 | Berkshire, Buckinghamshire, Gloucestershire, Wiltshire, Bristol, South Somerset, and Liverpool (urban and suburban) | Sub-sample 13-18 years | Short Child Maltreatment Questionnaire (excluding 1 item relating to sexual abuse) | RCADS-25 | - | 2 Self-report items.<br>1. Frequency of going to bed hungry<br>2. Degree of worry concerned with money to pay for food and living costs | - | University of Oxford Research Ethics Committee (ref: R62366) |
| REACH | Over 4,000 | London (urban) | 11-14 years | 4 Scales:<br>1. Self-report adolescent-appropriate Life Events Checklist<br>2. 9 items assessing school exclusions, receiving foster care and homelessness. | GAD-7, SMFQ | SDQ hyperactivity/inattention subscale | 6 Self-report items:<br>1. Free school meals<br>2. Number of rooms in house<br>3. Have your own bedroom<br>4. Family car ownership | Social Support:<br>1. Multidimensional Scale of Perceived Social Support (12 item)<br>2. Self-report items on help-seeking from professionals and | Psychiatry, Nursing and Midwifery Research Ethics Subcommittee, King's College London (ref:15/162320) |
|  |  |  |  | 3. Bullying victimisation<br>SDQ item<br>4. Revised Olweus<br>Bully/Victim<br>Questionnaire |  |  | 5. Ownership of<br>electronic devices<br>6. Been on holiday in<br>the past 12 months | perceived quality of<br>social relationships<br>Cognitive factors:<br>1. CCSC<br>2. CASQ-R<br>3. CAMM |  |
| DASH | Over<br>6,500 | London (urban) | 11-23<br>years | Self-report items<br>covering<br>parental death, mental<br>and physical illness,<br>foster care, separation<br>from parents,<br>harassment and<br>discrimination due to<br>gender, race, religion or<br>other personal<br>characteristics (e.g.,<br>physical appearance). | SDQ emotional<br>problems<br>subscale | SDQ<br>hyperactivity/<br>inattention<br>subscale | A total disadvantage<br>score – calculated by<br>the sum of<br>endorsement of 37<br>items relating to<br>access to household<br>provisions | Social support:<br>1. Multidimensional<br>Scale of Perceived<br>Social Support<br>2. Self-report items | The Multicentre<br>Research Ethics<br>Committee and<br>NHS Local<br>Research Ethics<br>Committees. |

*Abbreviations:* SDQ, Strengths and Difficulties Questionnaire; RCADS-25, Revised Child Depression and Anxiety Scale-25; GAD-7, Generalized Anxiety Disorder Scale-7; SMFQ, Short version of the Moods and Feelings Questionnaire; CCSC, Children’s Coping Strategies Checklist; CASQ-R, Children’s Attributional Style Questionnaire – Revised; CAMM, Child and Adolescent Mindfulness Measure.

### Measures

Variables of interest for ATTUNE work package 2 were assessed using different instruments in each cohort and are summarised in ***Table 1*** and described in detail below.

### Adverse childhood experiences

<u>HeadStart Cornwall</u> measured adverse experiences in childhood and adolescence using two approaches. First, using a bullying victimisation self-report item from the Strengths and Difficulties Questionnaire [SDQ] [31]: “Other children or young people pick on me or bully me” which was rated on a 3-point Likert scale ranging from “not true” to “certainly true”. Second, using data collected from the local government Supporting Families programme, covering a range of experiences which would mean that a child or their family would engage with services. These include, risk of sexual exploitation, homelessness, exposure to domestic violence and unmanaged physical or mental illness in the household.

In <u>Oxwell,</u> experiences of youth adversity was indexed using 6 items from the Short Child Maltreatment Questionnaire, excluding 1 original item relating to sexual abuse [32]. The questionnaire measures 6 forms of maltreatment covering abuse, neglect and the witnessing of domestic violence. Respondents rate whether they have experienced each of these forms of maltreatment, and the frequency (once or twice, many times).

<u>REACH</u> assessed youth adversity using the 16-item self-report adolescent-appropriate Life Events Checklist [33]. Respondents rate whether they have experienced events including the death of someone close, being the victim of a crime, parental separation, and the experience of a serious accident or illness. A further 9 items were used to assess other forms of youth adversity including school exclusions, receiving foster care, and homelessness. The bullying victimisation SDQ item (as above) and the 4-item Revised Olweus Bully/Victim Questionnaire [34], were used to assess bullying victimisation.

In <u>DASH,</u> a series of self-report items covering a range of experiences and circumstances were used to measure youth adversity. These include parental death, mental and physical illness, foster care, separation from parents, harassment and discrimination due to gender, race, religion or other personal characteristics (e.g., physical appearance) [29].

### Depressive and anxiety symptoms

<u>HeadStart Cornwall and DASH</u>: the emotional problems subscale of the SDQ [31] was used to measure symptoms of depression and anxiety. This subscale consists of 5 items rated on 3-point Likert scale from “not true” to “certainly true”.

<u>OxWell</u> used the Revised Child Anxiety and Depression Scale (RCADS-25) [35]. The scale has 25 items, which are rated on a 4-point Likert response scale (never, sometimes, often, always).

<u>REACH</u> used 2 self-report instruments to measure depression and anxiety. The Short Moods and Feelings Questionnaire was used to assess core depressive symptoms using 13 self-report items rated on a 3-point Likert scale ranging from “not true” to “true” [36]. Anxiety was measured using the 7-item Generalised Anxiety Disorder Scale-7 [37]. Participants are required to rate the frequency of each of the symptoms ranging from “not at all” to “nearly every day”.

### ADHD traits as a measure of neurodivergence

<u>HeadStart Cornwall, REACH, and DASH</u>: The SDQ hyperactivity/inattention subscale [31] was used to assess innate neurodivergence (traits of ADHD). This subscale is comprised of 5 items rated on a 3-point Likert scale from “not true” to “certainly true”.

### Socio-economic status

In HeadStart Cornwall receipt of free school meals will be used as a proxy for SES. These data is drawn from official School Census records.

<u>OxWell</u> measured SES using 2 questions. The first question is: “Some young people go to school or to bed hungry because there is not enough food at home. How often does this happen to you?”, the frequency of which is rated using 4 options ranging from “not at all” to “everyday”. The second question is: “To what extent do you worry about having enough money to pay for food or living costs?”, rated on a 5-point Likert scale ranging from “not at all” to “extremely worried”.

<u>REACH</u> assessed SES using a series of self-report items covering free school meals, size of family home (number of bedrooms, participant having their own room), family car ownership, ownership of electronic devices (laptop, tablet), and holidays in the last year [28].

In <u>DASH</u>, a total disadvantage score was calculated using the sum of endorsement of 37 items relating to access to household provisions, for example, “Does your family have a garden?”, and “Do you have your own bedroom?”.

### Social support

<u>REACH and DASH</u> used the Multidimensional Scale of Perceived Social Support to assess social support [38]. This instrument consists of 12 items rated on a 5-point Likert scale (from “strongly disagree” to “strongly agree”) and covers support received from family, friends and significant others. DASH also used a series of self-report items to assess a range of experiences that could be classified as social support. These covered engagement with religious groups (e.g., church attendance), support from family, frequency of family activities, support/relationship with parents, engagement in recreational activities (e.g., sports). Additionally, REACH enquired about help-seeking from a range of professionals [39] and quality of social relationships using self-report items.

### Cognition

<u>REACH</u> used 3 instruments to measure aspects of social cognition. First is the Children’s Coping Strategies Checklist [CCSC] [40]. Twenty-six items were extracted from the original CCSC to assess four types of coping: distraction, support seeking, active, and avoidant. Each item was rated on a 4-point Likert scale ranging from “never” to “most of the time”.

Second, the Children’s Attributional Style Questionnaire-Revised (CASQ-R) containing 24 items to assess children’s attributional style [41]. The questionnaire consists of 12 positive (e.g., “you make a new friend”) and 12 negative events (e.g., “you break a glass”), each followed by 2 possible causes for the event, varying on one of three dimensions of attributional style (internal versus external, stable versus unstable, and global versus specific).

Third, the Child and Adolescent Mindfulness Measure (CAMM) containing 10 items to measure dimensions of mindfulness (e.g., “I push away thoughts that I don’t like”) [42]. Each item was rated on a 5-point Likert scale ranging from “never true” to “always true”.

### Ethical considerations

Participants from all cohorts provided consent (or assent and parental consent if aged 16 years or younger). Ethical approval was obtained for all cohorts from various Ethics committees across the UK, details of which are provided in ***Table 1***. Cohort data used for this current programme of research will be provided in an anonymised format.

### Statistical analyses

The analyses that will be used to address each of the research questions for RQ2-4 are outlined below, both from a structural equation modelling and a multilevel modelling approach, based on the available data. Note that different research questions will be addressed with different cohorts (see ***Table 1***). For RQ1, which is focused on differences across cohorts in the prevalence of youth adversity as well as average levels of depressive and anxiety symptoms, descriptive statistics will be presented for each cohort separately. Effect sizes of the differences between the prevalence (youth adversity) and means (depressive and anxiety symptoms) for the different levels of the demographic characteristics (e.g., low SES versus high SES) will be reported. Linear regressions of the effects of youth adversity and the demographic characteristics on depressive and anxiety symptoms will be conducted. Models will be compared across cohorts to assess differences and will be treated as emergent analysis. With follow up analysis conducted where needed, appropriate and possible.

### Structural equation modelling

In the following models, group will be defined by intersectionality profile (e.g., female, low SES and high ADHD traits). Clustering in the data will be represented by dummy variables, entered into the model as covariates and constrained to equality across groups.

RQ2: A multiple group model of depression and anxiety symptoms regressed on youth adversity will be conducted. Preceding these regression analyses, measurement invariance of depressive and anxiety symptoms will be assessed across intersectionality profiles.
RQ3: A conditional multiple group latent growth model of depressive and anxiety symptoms will be conducted, where the latent growth factors are regressed on youth adversity. Depressive and anxiety symptom total scores (at each time-point) will be treated as continuous data.
RQ4: Multiple group mediation analysis will be conducted, where the social support variables are regressed on youth adversity, and depressive and anxiety symptoms are regressed on youth adversity and social support.

### Multilevel modelling

Where there are 20 or more intersectionality profiles (e.g., female, low SES and high ADHD traits) a multilevel modelling framework will be used to address the research questions. Specifically, multilevel analysis of individual heterogeneity and discriminatory accuracy (MAIDHA) will be used to model intersectionality profiles at the strata (cluster) level [43]. In these analyses, intersectionality profiles will not be treated as a grouping variable but as a strata-level variable. Youth adversity, as well as the individual-level characteristics contributing to the intersectionality profiles will be entered as main effects, and residual-level variation will be considered to reflect the intersection of the individual-level characteristics.

### Inference criteria

Effect sizes will be reported, and *p-*values for estimated regression parameters will be corrected for multiple testing using the false discovery rate method, where appropriate [44] for comparing multiple conditions within the same hypothesis. The Bayesian information criterion (BIC) will be the primary criterion for comparing the fit of non-nested models, as well as the Akaike information criterion (AIC). For non-saturated models, the comparative fit index (CFI), standardised root mean square residual (SRMR), and root mean square error of approximation (RMSEA) values will be used to assess model fit. CFI > .95, SRMR <.06, and RMSEA <.08 will broadly be considered to indicate acceptable fit [45]. For multilevel models, the variance partition coefficient will be used to assess effects at the (strata) residual level of intersectionality.

### Missing data

Where dependent data is continuous, missing data will be assumed to be missing at random and will be accommodated using full information maximum likelihood estimation. Where dependent data is ordered categorical, pairwise present data will be used, with weighted least squares estimation. Listwise deletion will be applied for missing covariate data and will be accompanied by sensitivity analyses to explore the nature of the missing data.

### Status and timeline

The development work for this research programme is mature and almost complete. It covers statistical analysis plans (as outlined above) and mapping of papers to determine the publication strategy for the results. It is anticipated that the core data analysis will be undertaken and completed between 2023-2024. ATTUNE Young People’s Advisory Groups [YPAGS] have been consulted regarding the conceptualisation of variables to develop analysis plans and are booked at several points during that period. The goal will be to disseminate the findings as they become available (including scientific article publication) rather than solely at the end of this period. Pre-registration of the work will be submitted individually by research question/cohort.

## Discussion

The aim of Work Package 2 in the ATTUNE project is to explore the role of intersectionality in the relationship between youth adversity and mental health in young people, including the examination of social and cognitive mechanisms. To date, research has primarily focused on the variations between groups based on individual characteristics (e.g., gender or ethnicity), but less so on their intersections (e.g., gender and ethnicity) in relation to the prevalence of and the strength of association between youth adversity and mental health problems [8]. Focusing on intersectionality in ATTUNE Work Package 2 will provide new insights into specific groups that may benefit most from prevention and intervention efforts, potentially enhancing both the efficacy and efficiency of such work.

The planned research programme has several methodological strengths, including the large sample sizes and diverse geographical locations across the UK in the cohorts studied. However, there are also several methodological limitations that should be anticipated and considered when undertaking this work and interpreting the results. Firstly, the majority of the data that will be analysed are derived from self-report instruments. Although such instruments have been shown to be reliable sources of information, particularly in terms of assessing internal and mood states (e.g., depression and anxiety) [46], they are also associated with some biases, especially with regards to reporting experiences of youth adversity. For example, inaccuracies in the data obtained from self-report youth adversity questionnaires may arise due to normal forgetting, current mood, and factors such as infantile and traumatic amnesia [47, 48]. Nevertheless, self-report measures have been reported to exhibit very good reliability and validity, even in clinical samples [49]. Moreover, not all data that will be analysed as part of this research programme are subject to such biases, as some are drawn from official records of youth adversity [26].

Another key limitation that should be considered is the lack of consistency in assessments across the cohorts used here. Consequently, any observed differences in results may be due to variations in assessment methods employed by each cohort, rather than factors such as place or other factors of interest (e.g., different intersectionality profiles). Thus, caution must be taken when interpreting the findings.

### Youth involvement

Consultation with YPAGs convened by the wider ATTUNE project will be undertaken throughout the work package’s life cycle. Discussion and feedback with the YPAGS have already been provided regarding conceptualisation of youth adversity. Plans are in place to work with the YPAGS with regards to the interpretation of results for each of the research questions, and identification of social and cognitive mechanisms. Feedback on dissemination plans will also be sought from the YPAGS.

### Dissemination plans

The findings from this work package will be disseminated through multiple channels, specifically targeting a diverse range of stakeholders. The team will leverage the extensive network established for the ATTUNE project, which includes charities (focused on youth mental health and support for victims of youth adversity), policymakers, mental health practitioners, and professionals working with young people. A central aspect of ATTUNE is the involvement and representation of youth voices in all aspects of the project. Guidance on effectively reaching this diverse audience will be sought from the ATTUNE research collaboration network. Key communication channels for dissemination will include peer-reviewed journal articles, conference presentations, lay summaries, social media posts (e.g., twitter) and public engagement events.

### Summary

This extensive research programme will explore the role of intersectionality on the relationship and (social and cognitive) mechanisms between youth adversity and mental health problems in young people living across the UK. The anticipated novel findings will not only broaden our understanding of the influence of youth adversity on mental health outcomes in different groups, but will have important clinical implications, which may help identify those at greatest risk of poor mental health and thus, those who may benefit most from intervention efforts.

### Financial disclosures

The ATTUNE project is funded by a cross council UK Research and Innovation [UKRI] award (MR/W002183/1). The HeadStart Cornwall data was collected as part of the HeadStart learning programme and supported by funding from The National Lottery Community Fund. The content is solely the responsibility of the authors, and it does not reflect the views of The National Lottery Community Fund. We are grateful to all of the research teams and participants who have contributed to the data that will be analysed in this work package.

## Data Availability

No datasets were generated or analysed during the current study. The project will undertake secondary analysis of several existing datasets which does not belong to the research team

